# Malnutrition and healthcare costs in older adults in Sweden: a longitudinal study based on a population-based cohort and Swedish registers

**DOI:** 10.64898/2026.05.29.26354412

**Authors:** Xin Xia, Yohannes Mengistu Balcha, Adrián Carballo-Casla, Emil Aho, Carl Willers, Elisabeth Rydwik, Amaia Calderón-Larrañaga, Susanna Kugelberg, Aravinda Berggreen-Clausen, Josephine Garpsäter, Linus Jönsson

**Author notes:** **Corresponding author:** Xin Xia, **Address:** Karolinska Institutet, BioClinicum, Solnavägen 30, floor 10, 171 64 Solna, Sweden, **Email:**.

## Abstract

**Background:** The study aimed to estimate healthcare costs associated with malnutrition in Swedish older adults.

**Methods:** We conducted a cohort study using data from the population-based Swedish National Study on Aging and Care in Kungsholmen (SNAC-K, N = 2982), a geriatric inpatient cohort of complex patients (N = 7680), and a cohort of individuals with cognitive impairment from the Swedish Register of Cognitive/Dementia Disorders (SveDem, N = 64192). At risk of malnutrition and malnutrition were ascertained by the Mini-Nutritional Assessment in SNAC-K and the geriatric inpatient cohort. In SveDem, body mass index was used for identifying malnutrition. Healthcare resource use was derived from regional and national registers. Associations between malnutrition and healthcare costs in 2024 Swedish kronor (SEK) were analyzed using two-part models and generalized linear regression models, adjusting for demographic and clinical factors.

**Findings:** In the community, at risk of malnutrition and malnutrition were associated with an increase in annual healthcare costs of 2267 SEK (95% CI: 64,4469) and 1846 SEK (95% CI: - 6802,10493), respectively. In geriatric patients, healthcare costs over 6 months in individuals at risk of malnutrition and individuals with malnutrition were 60205 SEK (45613,74798) and 86619 SEK (68362,104875) higher than those without malnutrition. In people with cognitive impairment, malnutrition was associated with higher annual healthcare costs (22170 SEK, 95% CI: 15152,29188).

**Interpretation:** Both at risk of malnutrition and malnutrition are associated with higher healthcare costs in Swedish older adults. The study findings are important for informing future economic evaluations of malnutrition interventions in Swedish older adults.

**Funding:** The study received funding from a research agreement with the Swedish Food Agency (agreement number: 2025-03258).

## Introduction

Undernutrition, commonly referred to as malnutrition, is a condition defined as insufficient nutrient intake relative to the body’s needs and is prevalent among older adults [1, 2]. Many conditions common in older adults can contribute to malnutrition, including age-related physiological changes (e.g., age-related appetite loss), physical and mental impairments (e.g., multimorbidity), and social factors (e.g., social isolation) [2]. Consequently, the etiology of malnutrition in older adults is often multifactorial [1, 2]. Management is therefore recommended to be individualized, addressing not only inadequate nutrient intake but also the underlying causes of malnutrition [1, 2].

Malnutrition is a risk factor for various adverse health outcomes in older adults, including frailty, sarcopenia, falls, pressure ulcers, and longer inpatient stays [1, 2]. Malnutrition is also associated with increased healthcare costs in older adults, and studies from several developed countries have demonstrated its substantial economic consequences [3-7]. For example, at the individual level, a population-based cohort study in people aged 70 years and older in Spain found that annual healthcare costs were €714 higher in people with malnutrition than those without malnutrition [4]. At the national level, the estimated annual costs of disease-related malnutrition in older adults were €1.5 billion in the Netherlands and $4.3 billion in the United States [5, 6]. These studies provide valuable insights into the economic consequences of malnutrition in older adults on healthcare systems, which are important for informing economic evaluations of interventions. However, they have limitations, including a primary focus on hospital costs or disease-related malnutrition. Furthermore, malnutrition commonly affects older adults with dementia, a condition that incurs significant healthcare costs, but whether malnutrition further increases healthcare costs in this population has not been examined [8, 9].

Similar to other developed countries, Sweden has a growing older population and thus potentially a substantial number of individuals with malnutrition and associated healthcare costs. Sweden has well-established healthcare registers and population-based research cohorts of older adults, enabling comprehensive evaluation of the costs of malnutrition across different populations and types of healthcare resources [10-12]. Despite the potential economic burden of malnutrition and the availability of such data sources, no study has systematically assessed the healthcare costs of malnutrition among older adults in Sweden.

Therefore, in this study, we sought to use data from a population-based cohort and Swedish registers to assess healthcare costs associated with malnutrition among the general older adult population, as well as among older adults with functional impairment and complex health conditions and older adults with cognitive impairment.

## Methods

### Study design and study populations

The study consisted of three study populations derived from a Swedish population-based cohort and several Swedish registers with different observational periods (**Figure 1**). The three study populations aimed to cover three target populations in Sweden: older adults in general, older adults with functional impairment and complex health conditions, and older adults with cognitive impairment.

**Figure 1.**
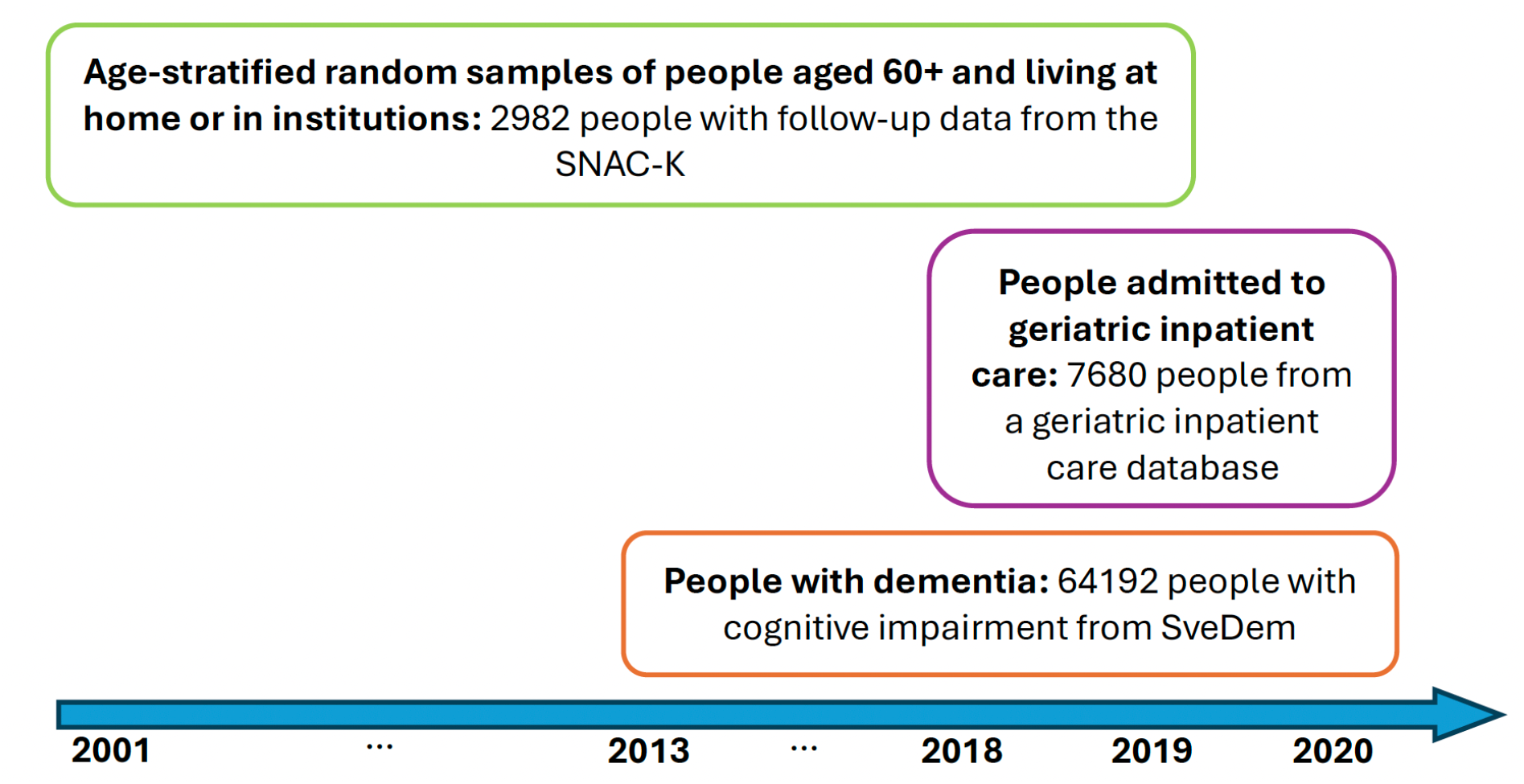
Study populations and observational periods. Abbreviations: SNAC-K = The Swedish National Study on Aging and Care in Kungsholmen, SveDem = the Swedish Register of Cognitive/Dementia Disorders.

The first study population included 2982 people with follow-up data from the Swedish National Study on Aging and Care in Kungsholmen (SNAC-K), an ongoing population-based cohort study. The SNAC-K recruited participants aged 60+ living at home or institutions in Kungsholmen, the central area of Stockholm, using an age-stratified random sampling strategy [10]. The baseline evaluation in the SNAC-K occurred between 2001 and 2004, after which people aged <78 years and people aged 78+ years at baseline have been followed every six years and every three years, respectively. In the present study, we used follow-up data from SNAC-K until 2019. The second study population consisted of 7680 people from a register of individuals admitted to geriatric inpatient care between 2018 and 2019 in Stockholm, which mainly covers older adults with functional impairment or with multiple serious medical conditions [11]. Six-month follow-up data of these individuals were available for the current study. The third study population included 64193 individuals diagnosed with dementia or the prodromal stage of dementia between 2013 and 2020 in Sweden and was derived from the Swedish Register of Cognitive/Dementia Disorders (SveDem) [12]. The study population was followed until the end of 2020. SveDem is a national register of people with dementia diagnosed at primary and specialist care and covers all memory clinics and 78% of primary care units in Sweden [12].

The SNAC-K was approved by the Regional Ethical Review Board in Stockholm, and the current study is within the scope of the ethical approvals. Written informed consents were obtained from all participants or their next of kin in the SNAC-K. Ethical approvals were obtained from the Swedish Ethical Review Authority for the use of data from the geriatric inpatient cohort and SveDem in the present study (DNRs: 2025-04479-02, 2024-07272-02). Written informed consents are not required for register-based research in Sweden.

### Measurements of malnutrition, healthcare costs, and covariates

#### The SNAC-K cohort

Malnutrition was ascertained using the Mini-Nutritional Assessment (MNA) scale, which categorized individuals into normal nutritional status, at risk of malnutrition, and malnourished [13]. The items included in MNA, their scoring, and deviations in the SNAC-K cohort are reported in **Supplementary Methods**.

Data on primary care were obtained from the healthcare administrative register in Stockholm (VAL database) and were available until the end of 2011 in the SNAC-K cohort [14]. Data on specialist outpatient and inpatient care were obtained from the Swedish National Patient Register (NPR) and were available throughout the observational period [15]. Death was ascertained by the Swedish Cause of Death Register (CDR) [16]. To calculate costs of healthcare, we used data on average per-visit costs for hospitals and clinics obtained from the cost per patient (KPP) database (**Supplementary Methods**) [17].

We considered age, sex, social isolation, and chronic diseases as key confounders in the association between malnutrition and healthcare costs. Living alone was used as a proxy for social isolation and was ascertained through interviews by SNAC-K nurses. Chronic diseases at baseline were ascertained by combining information from medical history and interviews conducted by SNAC-K physicians and data from VAL and NPR using the International Classification of Diseases (ICD) codes (**Supplementary Methods**). Chronic diseases considered in this study included ischemic heart disease, heart failure, stroke, Parkinson’s disease, dementia, chronic obstructive pulmonary disease, rheumatoid arthritis, osteoporosis, cancer, and depression. These chronic diseases were chosen because they are strong risk factors for malnutrition in older adults [2]. Other potential confounders included educational level and smoking status, which were ascertained through interviews by SNAC-K nurses.

#### The geriatric inpatient care cohort

Malnutrition was evaluated using the MNA scale in the geriatric inpatient care cohort, which did not deviate from the MNA.

The geriatric inpatient care cohort derived information on inpatient and outpatient care six months before and after admissions to geriatric inpatient care from the VAL database. Primary care costs were calculated using average costs per type of visit, and specialist inpatient and outpatient care costs were calculated using costs per diagnosis-related group (DRG) code from the KPP database (**Supplementary Methods**). Social care information was obtained from the Swedish National Register of Care and Social Services for the Elderly and Persons with Impairments (SOL), and social care costs were estimated using unit costs from the municipality and region database (kolada.se) and the organization that calculated the unit costs for the database (**Supplementary Methods**) [18]. Death was ascertained from population databases at Statistics Sweden.

We considered the same set of key confounders in this cohort and additionally considered disability level. Living alone was ascertained from population databases at Statistics Sweden. Disability level was measured using the Barthel index [11]. Chronic diseases before admissions to geriatric inpatient care were ascertained based on records in VAL and medical records at geriatric inpatient admission using ICD-10 codes (**Supplementary Methods**).

Other potential confounders included educational level and civil status, which were obtained from population databases at Statistics Sweden.

#### The SveDem cohort

There was no nutritional assessment in SveDem. Therefore, we used body mass index (BMI) information as a proxy for nutritional assessment and defined malnutrition as BMI <20 kg/m^2^ for individuals aged < 70 years and BMI <22 kg/m^2^ for individuals aged 70+ years [19].

Data on specialist outpatient care and inpatient care were obtained from the NPR, and their costs were estimated using costs per DRG from the KPP database (**Supplementary Methods**) [17]. Drug prescription cost data were obtained from the National Prescribed Drug Register (PDR) [20]. Social care costs were estimated through the same approach as in the geriatric inpatient care cohort (**Supplementary Methods**). Death was ascertained from the CDR.

Chronic diseases were ascertained from the NPR using ICD-10 codes and complemented by information on drug prescription from the PDR using Anatomical Therapeutic Chemical Classification System (ATC) codes to reduce the limitation of not having primary care information (**Supplementary Methods**). Dementia was not considered a separate chronic disease in this cohort, as everyone was diagnosed with dementia or the prodromal phase of dementia. Other covariates considered as potential confounders for the association between malnutrition and healthcare costs included dementia subtypes, cognitive impairment level, and institutionalization status, which were ascertained from SveDem.

Healthcare costs were inflated to 2024 Swedish kronor (SEK) using the Consumer Price Index from Statistics Sweden for all three cohorts [21].

### Statistical analysis

Overall prevalence and prevalence by age groups of malnutrition were estimated. Age groups were defined differently in the SNAC-K (60-72 years, 78+ years) than the geriatric clinic cohort and the SveDem cohort (<70 years, 70-79 years, and 80+ years), because SNAC-K used two pre-defined age cohorts that determined different follow-up schemes.

The associations between malnutrition and healthcare costs were evaluated using two-part models, which allowed for the modeling of two data-generating processes: one governing the probability of incurring healthcare costs (binary outcome) and the other governing the amount of healthcare costs (continuous outcome) among those who incurred costs [22]. Logistic regression was chosen for the binary outcome component. For the continuous outcome component, generalized linear regressions with different link functions and distributions were fitted, and the final models were determined based on model fitness measures and differed across the three cohorts. An exception was made when evaluating the associations between malnutrition and total costs and geriatric inpatient care costs in the inpatient geriatric cohort, for which generalized linear regression models were used instead of two-part models. This approach was chosen because admission to geriatric inpatient care inherently indicated healthcare utilization, eliminating the need to model the probability of incurring healthcare costs. The two-part models and the generalized linear regression models were adjusted for age, sex, follow-up year, and death during the year, and then additionally adjusted for living alone, chronic diseases, and cohort-specific covariates, depending on the data availability.

Marginal means of annual healthcare costs by nutritional status, as well as marginal effects of malnutrition on annual healthcare costs, were predicted. Marginal means of annual healthcare costs for the no malnutrition group were estimated by setting covariates to their observed values and nutritional status to no malnutrition for each individual, then averaging the predicted values. Marginal means for the at risk of malnutrition and malnutrition groups were estimated using the same procedure. The marginal effect of malnutrition was estimated as follows. For each individual, annual healthcare costs under each nutritional status were predicted by setting covariates to the individual’s observed values and varying nutritional status. The individual-level marginal effect was calculated as the difference in predicted annual healthcare costs between nutritional status categories. The marginal effects across all individuals were then averaged to obtain the overall marginal effect of malnutrition. In the geriatric inpatient care cohort, only 6-month care utilization data after admission were available; thus, marginal means of six-month healthcare costs were predicted.

We also estimated the costs of malnutrition in Sweden in 2024 at the national level by combining marginal effects of malnutrition on annual healthcare costs with population statistics. These statistics comprised the number of inhabitants and the number of individuals admitted to geriatric inpatient care in Sweden in 2024, categorized into 5-year age intervals (**Supplementary Methods**). The costs of malnutrition in Sweden were estimated by multiplying the number of inhabitants by the marginal effects of malnutrition on annual healthcare costs and subsequently summing the resulting products.

Missing data on malnutrition were handled using multiple imputation by chained equations in all analyses except for the analyses for describing the study populations. Data preparation was done using R (version 4.5.2). Statistical analyses were conducted using Stata version 19.5 (StataCorp LLC). The two-part model analyses were performed using the user-written Stata package “twopm” [22]. Marginal means and marginal effects were estimated using the Stata command “margins”.

## Results

The average age of the SNAC-K cohort (75.0 years) was lower than that of the geriatric inpatient care cohort (83.1 years) and the SveDem cohort (80.2 years), and there were more females than males in all three cohorts (**Table 1**). The proportion of people living alone was higher in the SNAC-K cohort and the geriatric inpatient care cohort than in the SveDem cohort. Only a small proportion of people in the SNAC-K cohort and the geriatric inpatient care cohort lacked malnutrition data, while more than 30% of individuals in the SveDem cohort did not have malnutrition data. The burden of chronic diseases differed across the three study populations. For instance, heart failure and chronic obstructive pulmonary disease were more prevalent in the geriatric inpatient care cohort, whereas stroke and depression were more prevalent in the SveDem cohort compared with the other cohorts. More details of the characteristics of the study populations are reported in **Table S1, Table S2**, and **Table S3**.

**Table 1.** Baseline characteristics of the study populations by cohort.

|  | **SNAC-K**<br>**(N = 2982)** | **Geriatric inpatient care**<br>**cohort (N = 7680)** | **SveDem**<br>**(N = 64192)** |
| --- | --- | --- | --- |
| **Age (years), mean (SD)** | 75.0 (11.3) | 83.1 (8.4) | 80.2 (7.9) |
| **Sex – female, n (%)** | 1937 (65.0) | 4693 (61.1) | 37029 (57.7) |
| **Living alone, n (%)\*** | 1755 (58.9) | 4509 (58.7) | 28118 (43.8) |
| **Malnutrition status, n (%)\*** |  |  |  |
| Normal nutritional status | 1918 (64.3) | 1478 (19.2) | 33910 (52.8) |
| At risk of malnutrition | 781 (26.2) | 3914 (51.0) |  |
| Malnourished | 92 (3.1) | 1796 (23.4) | 10435 (16.3) |
| **Chronic diseases, n (%)** |  |  |  |
| Ischemic heart disease | 514 (17.2) | 1340 (17.4) | 10842 (16.9) |
| Heart failure | 354 (11.9) | 2015 (26.2) | 7253 (11.3) |
| Stroke | 305 (10.2) | 646 (8.4) | 9451 (14.7) |
| Parkinson's disease | 31 (1.0) | 261 (3.4) | 2648 (4.1) |
| Chronic obstructive pulmonary disease | 157 (5.3) | 1186 (15.4) | 3272 (5.1) |
| Rheumatoid arthritis | 59 (2.0) | 156 (2.0) | 1003 (1.6) |
| Osteoporosis | 232 (7.8) | 938 (12.2) | 3273 (5.1) |
| Cancer | 603 (20.2) | 1191 (15.5) | 10669 (16.6) |
| Depression | 316 (10.6) | 551 (7.2) | 18216 (28.4) |
| Dementia | 164 (5.5) | 1000 (13.0) | - |

Abbreviations: SD = standard deviation, SNAC-K = The Swedish National Study on Aging and Care in Kungsholmen, SveDem = the Swedish Register of Cognitive/Dementia Disorders.
Notes: \*The numbers of people without data on living alone were 42 (0.5%) in the geriatric inpatient care cohort and 3883 (6.0%) in the SveDem cohort. The number of people without data on nutritional status was 191 (6.4%) in the SNAC-K, 492 (6.4%) in the geriatric inpatient care cohort, and 19847 (30.9%) in the SveDem cohort.

The estimated prevalence of malnutrition was higher among geriatric inpatient care patients (25.0%) and individuals with dementia (24.0%) than among the general older adult population (3.7%; **Table 2**). The prevalence of being at risk of malnutrition and the prevalence of malnutrition were higher in older age groups than in younger age groups in the general older adult population and in people with dementia, but not in geriatric inpatient care patients.

**Table 2.**
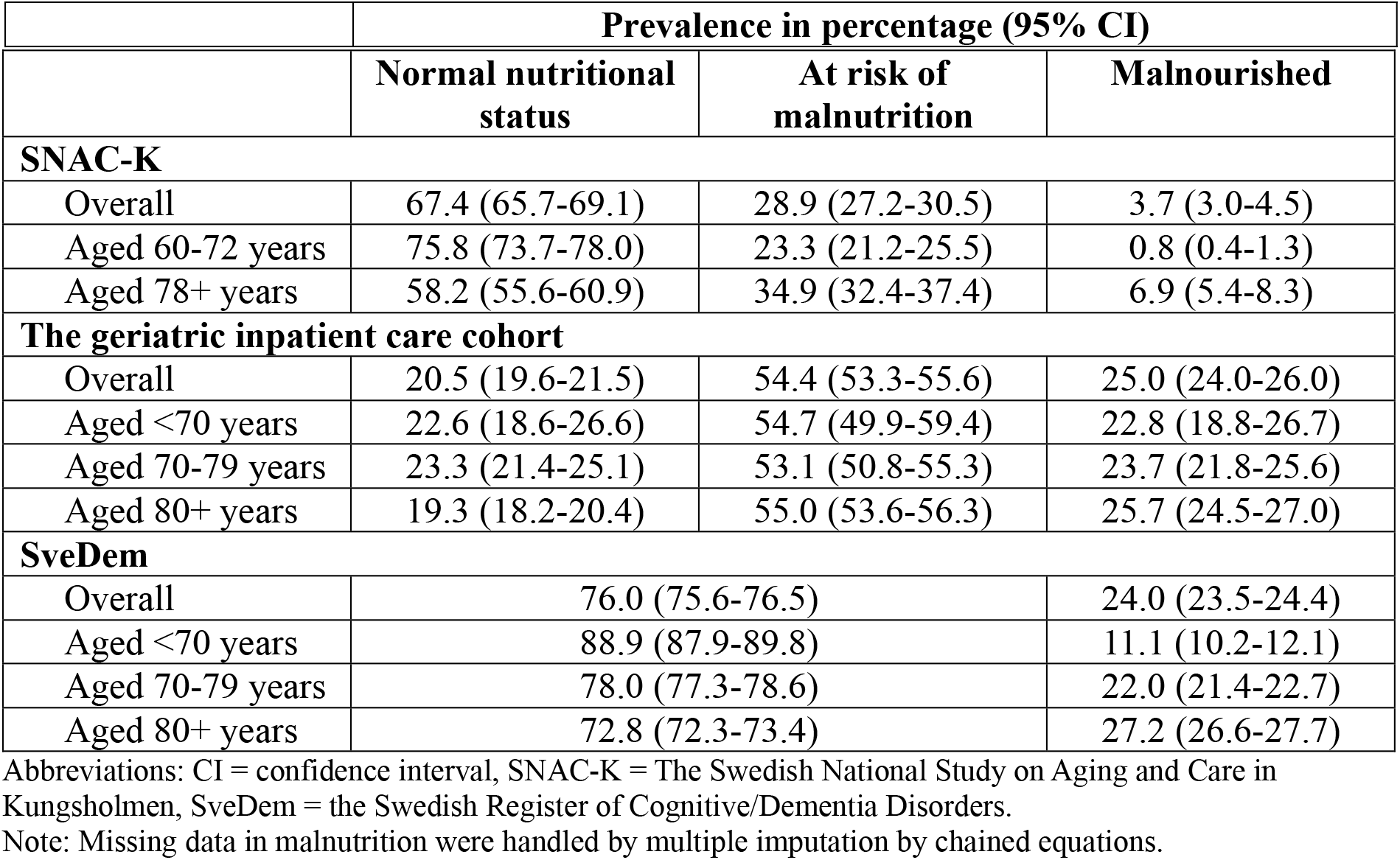
Overall prevalence and prevalence by age groups of malnutrition by cohort.

Combining the estimated prevalences of malnutrition with the population statistics in Sweden, the number of older adults at risk of malnutrition and the number of older adults with malnutrition in Sweden in 2024 were approximately 800000 and 102000, respectively.

On average, annual costs of healthcare were 2267 SEK (95% CI: 64,4469) higher for people at risk of malnutrition than for people with normal nutritional status in the general older adult population (**Table 3** and **Table S4**). Annual healthcare costs were also higher in people who were malnourished than those with normal nutritional status, but the difference (1846 SEK; 95% CI: -6802,10493) was not statistically significant, likely due to the small sample size of malnourished individuals. The differences in annual healthcare costs by nutritional status were largely attenuated after accounting for smoking status, living alone, and chronic diseases (**Table 3** and **Table S4**).

**Table 3.** Costs of healthcare in 2024 SEK by nutritional status.

|  | Model 1 |  | Model 2 |  |
| --- | --- | --- | --- | --- |
|  | Costs<br>(95% CI) | Difference<br>(95% CI) | Costs<br>(95% CI) | Difference<br>(95% CI) |
| **Annual costs of healthcare by nutritional status in the SNAC-K<sup>a</sup>** |  |  |  |  |
| Normal | 26724<br>(25539,27909) | Reference | 27575<br>(26365,28784) | Reference |
| At risk | 31706<br>(29473,33940) | 4982<br>(2543,7421) | 29841<br>(27841,31841) | 2267 (64,4469) |
| Malnourished | 33121<br>(25439,40803) | 6397 (-<br>1391,14185) | 29420<br>(20985,37855) | 1846 (-<br>6802,10493) |
| **Six-month costs of healthcare by nutritional status in the geriatric inpatient care cohort<sup>b</sup>** |  |  |  |  |
| Normal | 313332<br>(302259,324404) | Reference | 341558<br>(329385,353730) | Reference |
| At risk | 399471<br>(391061,407881) | 86139<br>(72066,100212) | 401763<br>(393335,410191) | 60205<br>(45613,74798) |
| Malnourished | 458522<br>(444299,472744) | 145190<br>(127056,163324) | 428176<br>(414979,441373) | 86619<br>(68362,104875) |
| **Annual costs of healthcare by nutritional status in SveDem<sup>c</sup>** |  |  |  |  |
| Normal | 501084<br>(497927,504242) | Reference | 505276<br>(502354,508199) | Reference |
| Malnourished | 541513<br>(535144,547883) | 40429<br>(32952,47906) | 527446<br>(521489,533404) | 22170<br>(15152,29188) |

Abbreviations: CI = confidence interval, SEK = Swedish kronor, SNAC-K = The Swedish National Study on Aging and Care in Kungsholmen, SveDem = the Swedish Register of Cognitive/Dementia Disorders.
Notes:
<sup>a</sup>Results are from marginal predictions from two-part models. Model 1 adjusted for age, sex, follow-up year, and death during the year. Model 2 additionally adjusted for smoking status, living alone, ischemic heart disease, heart failure, rheumatoid arthritis or osteoporosis, cancer, depression, and Parkinson's disease or dementia.
<sup>b</sup>Results are from marginal predictions from generalized linear regression models. Model 1 adjusted for age and sex. Model 2 additionally adjusted for educational level, civil status, living alone, functional ability, heart failure, depression, Parkinson's disease, and dementia.
<sup>c</sup>Results are from marginal predictions from two-part models. Model 1 adjusted for age, sex, dementia subtypes, follow-up year, and death during the year. Model 2 additionally adjusted for institutionalization status, living alone, severity of cognitive impairment, ischemic heart disease, heart failure, stroke, chronic obstructive pulmonary disease, rheumatoid arthritis, osteoporosis, and cancer.

In geriatric inpatient care patients, both being at risk of malnutrition (60205 SEK; 95%CI: 45613,74798) and being malnourished (86619 SEK; 95% CI: 68362,104875) were significantly associated with higher costs of healthcare six months after admission to geriatric inpatient care (**Table 3** and **Table S5**). The increased costs were driven by not only medical care but also social care (**Table S5**). The differences in costs of healthcare across nutritional status remained significant after accounting for functional ability and chronic diseases (**Table 3** and **Table S5**).

Among people with dementia, being malnourished was associated with a 22170 SEK (95% CI: 15152,29188) higher annual costs of healthcare, and the association remained evident after accounting for the level of cognitive impairment and chronic diseases (**Table 3** and **Table S6**). Notably, the difference was mainly driven by increased social care costs (**Table S6**).

At the national level, the estimated costs associated with being at risk of malnutrition in Swedish older adults in 2024 were approximately 4.9 billion SEK, while the costs associated with malnutrition were approximately 2.2 billion SEK (**Table S7**).

## Discussion

Using data from a Swedish population-based cohort and Swedish registers, the study found that not only malnutrition, but also being at risk of malnutrition, was associated with increased healthcare costs. The increase was observed among older adults in general, geriatric inpatients, and older adults with cognitive impairment, with the largest increase observed among geriatric inpatients. At the national level, the costs of healthcare associated with malnutrition in Sweden in 2024 were estimated at 2.2 billion SEK and 4.9 billion SEK for being at risk of malnutrition.

Estimating costs associated with malnutrition in general older adults is important for informing future economic evaluations of population-level interventions aimed at preventing or managing malnutrition as well as for giving an overview of the economic consequences of malnutrition at the national level. Our study provided the first estimates of healthcare costs associated with malnutrition in older adults in Sweden. In addition, it contributed new insights into the economic consequences of malnutrition among older adults. Previous research on malnutrition-related healthcare costs and healthcare resource utilization has typically focused on a single population, most often geriatric inpatients [4, 23-28]. The findings of the present study add to the literature that malnutrition was associated with increased costs of healthcare in older adults in general and older adults with cognitive impairment. Malnutrition in individuals with dementia is a distinct clinical concern, as all stages of dementia can affect nutrient intake through a range of neurological and behavioral mechanisms, such as loss of smell in early stages and forgetting to eat in later stages [8]. Furthermore, earlier studies were frequently limited to medical care costs, such as those associated with prolonged inpatient stays [4, 7]. The findings of the present study suggest that the economic consequences of malnutrition in older adults may extend beyond increased medical care costs (e.g., specialist inpatient care). Our study showed that, in geriatric inpatients and individuals with cognitive impairment, social care costs were also higher in individuals with malnutrition and in individuals at risk of malnutrition than in those with normal nutritional status, even after considering existing chronic diseases and functional ability. This finding is in line with previous research that showed malnutrition in older adults was associated with future functional decline, which may then lead to increased need for social care [29, 30].

The major strength of the study is that it leveraged longitudinal data from a population-based cohort and Swedish registers containing detailed information on healthcare resource use and potential confounders and included three groups of older adults. The individual-level data analyses provided greater flexibility in controlling for potential confounders, in contrast to previous research that relied on aggregated data on medical costs attributable to malnutrition for a limited number of diseases [5, 6]. In addition, the study did not restrict healthcare resource use to medical care but considered both medical care and social care. However, the study has several limitations. First, social care costs were not estimated for SNAC-K, which was included to represent the general older adult population, due to insufficient data.

Therefore, the healthcare costs associated with malnutrition in general older adults were likely underestimated. In addition, participants in SNAC-K had better socioeconomic status and better health than the general Swedish older adult population. This may have further contributed to an underestimation of healthcare costs associated with malnutrition. Second, the number of individuals with malnutrition in the SNAC-K was small, which may have resulted in limited statistical power to detect differences in healthcare costs between individuals with and without malnutrition. Third, malnutrition among individuals with cognitive impairment was not assessed using specialized nutritional assessment tools and was instead proxied by BMI. Consequently, the costs of being at risk of malnutrition in people with dementia could not be estimated. Future research in individuals with dementia that uses standard assessment tools to measure malnutrition status is warranted. Fourth, the temporality of chronic diseases, functional ability, and level of cognitive impairment relative to malnutrition could not be established. This may have led to overadjustment for mediators in the models, given that malnutrition can contribute to the development of these chronic diseases, functional decline, and cognitive decline. Lastly, only 6-month healthcare costs were analyzed for geriatric inpatients because longer follow-up data were unavailable. Therefore, our estimates of the national-level healthcare costs of malnutrition in 2024 relied on extrapolating 6-month costs to annual costs.

In conclusion, our study suggests that being at risk of malnutrition and malnutrition were both associated with medical and social care costs in older adults in Sweden. The increased costs of healthcare associated with malnutrition exist in general older adults, geriatric inpatients, and older adults with cognitive impairment. The findings provide important evidence on the economic consequences of malnutrition in older adults and have implications for informing interventions aimed at preventing and managing malnutrition across different older populations.

## Data Availability

We are unable to share the raw data used in this study due to Swedish legal restrictions. However, data from SNAC-K, the geriatric inpatient care cohort, and SveDem can be accessed following ethical approval and approval from the respective study steering groups.

## Contributors

LJ and XX conceptualized the study. XX and YMB analyzed the data. ACC, CW, ER, ACL, EA, XX, and LJ obtained the data for the study. XX drafted the manuscript of the study. All authors reviewed the manuscript, provided critical feedback, and approved the submission of the manuscript.

## Declaration of interests

The authors reported no conflicts of interest related to this study.

## Acknowledgments

Data collection of the Swedish National study on Aging and Care (SNAC-K) was supported by the Swedish Research Council (ongoing/current grant: 2021-00178); the Swedish Ministry of Health and Social Affairs; the participating County Councils and Municipalities. SveDem is funded by the Swedish Association of Local Authorities and Regions. The study received funding from a research agreement with the Swedish Food Agency (agreement number: 2025-03258).

XX received funding from Alzheimerfonden (AF-1033303). LJ is funded by the Innovative Health Initiative Joint Undertaking (101112145), PREDEM (Vinnova grant 2021-02680), the Swedish Research Council for Health, Working Life and Welfare, and Novo Nordisk. ER and CW is funded by Forte (2021-01793, 2024-01812). ER, ACL and LJ receive funding from the FORTE research center grant [grant number 2025-02071—”KI Transdisciplinary Research Center for Personalized Dementia Prevention & Care” (TraCeDem)].

Funding received by individual researchers is not related to the current work and had no impact on its design or execution.

## Notes

### Competing Interest Statement

The authors have declared no competing interest.

