## Supplementary methods for "Malnutrition and healthcare costs in older adults in Sweden: a longitudinal study based on a population-based cohort and Swedish registers"

### 1. Operationalization of nutritional status defined by Mini-Nutritional Assessment in the SNAC-K cohort

The Mini Nutritional Assessment (MNA) was not formally administered in the SNAC-K cohort. However, information on diet and other components included in the MNA was collected, allowing for the operationalization of nutritional status defined by MNA. The MNA criteria and deviations in their operationalization within the SNAC-K cohort are shown in **Table 1**.

**Table 1. Mini-Nutritional Assessment criteria and modifications in the SNAC-K cohort**

| Criteria | Score | Modifications in the SNAC-K cohort |
|---|---|---|
| Has food intake declined over the past 3 months due to loss of appetite, digestive problems, chewing or swallowing difficulties? | 0 = severe decrease in food intake<br>1 = moderate decrease in food intake<br>2 = no decrease in food intake | 0 = no appetite; food is tasteless; need to force oneself to eat<br>1 = slightly reduced appetite<br>2 = normal or increased appetite |
| Weight loss during the last 3 months | 0 = weight loss greater than 3 kg (6.6 lbs)<br>1 = does not know<br>2 = weight loss between 1 and 3 kg (2.2 and 6.6 lbs)<br>3 = no weight loss | 0 = weight loss greater than 3 kg (6.6 lbs)<br>1 = does not know; bothered by weight loss<br>2 = weight loss between 1 and 3 kg (2.2 and 6.6 lbs)<br>3 = no weight loss |
| Mobility | 0 = bed or chair bound<br>1 = able to get out of bed / chair but does not go out<br>2 = goes out | 0 = unable to independently get in and out of bed to a chair<br>1 = can move around indoors on their own or with assistance<br>2 = can move around outdoors on their own or with assistance |
| Has suffered psychological stress or acute disease in the past 3 months? | 0 = yes<br>2 = no | 0 = inpatient admission in the past three months; any of the following events in the past year: death of mother, death of father, parents' divorce or separation, physical abuse, divorce, death of spouse, loss of child, death of closest friend, grave illness, grave illness of close relative, moving from own home to specialized housing facility , become unemployed, |

| | | |
|---|---|---|
| | | retirement, severe financial loss, legal issues/trouble with justice<br>2 = none of the above |
| Neuropsychological problems | 0 = severe dementia or depression<br>1 = mild dementia or depression<br>2 = no psychological problems | 0 = dementia or depression<br>1 = cognitive impairment no dementia, mini mental state examination <24<br>2 = none of the above |
| BMI (kg/m$^2$) | 0 = BMI less than 19; calf circumference less than 31<br>1 = BMI 19 to less than 21<br>2 = BMI 21 to less than 23<br>3 = BMI 23 or greater; calf circumference 31 or greater | 0 = BMI less than 19; calf circumference less than 31<br>1 = BMI 19 to less than 21<br>2 = BMI 21 to less than 23<br>3 = BMI 23 or greater; calf circumference 31 or greater |

Abbreviations: BMI = body mass index, SNAC-K = the Swedish National Study on Aging and Care in Kungsholmen.
Note: Normal nutritional status is defined as scoring 12-14 points; at risk of malnutrition is defined as scoring 8-11 points; malnourished is defined as scoring 0-7 points.

### 2. Data sources for healthcare resource use and unit costs

Each cohort was linked to different data sources for healthcare resource use, which determined the cost components included in the analyses of each cohort in the present study. For example, the SNAC-K began in 2001, whereas the PDR was established in 2007. In addition, the SNAC-K was not linked to SOL. Consequently, drug costs and social care costs were not included in the analyses for costs of malnutrition in the SNAC-K cohort. The data sources for healthcare resource use and their availability in the three cohorts in the current study are presented in **Table 2**.

**Table 2. Data sources for healthcare resources**

| Data sources | Healthcare resources | Availability in the cohorts in the study |
|---|---|---|
| The health care administrative register in Stockholm (VAL database, held by Region Stockholm) [1] | All inpatient care and outpatient care (including primary outpatient care and specialist outpatient care) contacts in Stockholm, starting from the 1990s onward | • The SNAC-K<br>• The geriatric inpatient care cohort |
| The Swedish National Patient Register (NPR, held by Socialstyrelsen) [2] | • Nationwide inpatient care since 1987<br>• Specialist outpatient care since 2001<br>• Compulsory psychiatric care since 2010 | • The SNAC-K<br>• SveDem |
| The National Prescribed Drug Register (PDR, held by Socialstyrelsen) [3] | Prescribed drugs dispensed at pharmacies, starting from 2005 onward | • The SNAC-K<br>• SveDem |
| The Swedish National Register of Care and Social Services for the Elderly and Persons with Impairments (SOL, held by Socialstyrelsen) [4] | Social services, such as home care, institutional care, short-term residence, housing support, security alarm, meal delivery, and daytime activity, starting from 2007 onward | • The geriatric inpatient care cohort<br>• SveDem |

Abbreviations: SNAC-K = The Swedish National Study on Aging and Care in Kungsholmen, SveDem = the Swedish Register of Cognitive/Dementia Disorders.
Note: Socialstyrelsen is the Swedish National Board of Health and Welfare.

The data sources used to derive unit costs for healthcare resources differed between the SNAC-K and the geriatric inpatient care cohort because the available data on healthcare resource use also differed between the two cohorts (**Table 3**). In the geriatric inpatient care cohort, primary care costs were calculated using average costs per type of visit (e.g., per onsite visit to a primary care physician) from the cost per patient (KPP) database [5]. In the SNAC-K, the VAL database, which captured primary care contacts, was only available for the period 2001-2011, whereas information on the main caregiver was available from 2014 onward. Therefore, average costs by Swedish healthcare classification systems (MVO) codes (i.e., types of clinics) from the KPP database were used instead. In addition, diagnosis-related group (DRG) codes were not requested from the Swedish National Patient Register in the SNAC-K. Consequently, average costs by MVO codes were used to estimate costs of specialist care.

Unit costs for social care services in 2024 SEK were derived from the Kolada database and supplemented with data from the organization responsible for their calculation. This was necessary because Kolada does not consistently report all required unit costs.

**Table 3. Data sources for unit costs**

<table>
<tr><th></th><th>SNAC-K</th><th>The geriatric inpatient care cohort</th><th>SveDem</th></tr>
<tr><td>Primary care</td><td>Average costs by MVO codes from KPP database</td><td>Average costs per type of visit from KPP database</td><td>Not accounted</td></tr>
<tr><td>Specialist outpatient care</td><td>Average costs by MVO codes from KPP database</td><td>Costs per DRG code from KPP database</td><td>Costs per DRG code from KPP database</td></tr>
<tr><td>Specialist inpatient care</td><td>Average costs by MVO codes from KPP database</td><td>Costs per DRG code from KPP database</td><td>Costs per DRG code from KPP database</td></tr>
<tr><td>Drug prescription</td><td>Not accounted</td><td>Not accounted</td><td>PDR</td></tr>
<tr><td colspan="4">Social care</td></tr>
<tr><td>Housing support</td><td>Not accounted</td><td>Not accounted</td><td>269 SEK per day, Ensolution</td></tr>
<tr><td>Day activity</td><td>Not accounted</td><td colspan="2">148400 SEK per year, Kolada</td></tr>
<tr><td>Home care</td><td>Not accounted</td><td colspan="2">646 SEK per hour, Ensolution</td></tr>
<tr><td>Short-term residence</td><td>Not accounted</td><td colspan="2">3387 SEK per day, Ensolution</td></tr>
<tr><td>Institutionalization</td><td>Not accounted</td><td colspan="2">2566 SEK per day, Ensolution</td></tr>
</table>

Abbreviations: DRG = diagnosis-related group, Kolada = the municipality and region database, KPP = the cost per patient database, MVO = Swedish healthcare classification systems (Medicinska verksamhetsområden), PDR = the Prescribed Drug Register, SNAC-K = The Swedish National Study on Aging and Care in Kungsholmen, SveDem = the Swedish Register of Cognitive/Dementia Disorders.

### 3. Identification of chronic diseases using ICD codes and ATC codes

In the SNAC-K, 4-digit ICD-9 and ICD-10 codes were used for identifying chronic diseases, as data from the Swedish National Patient Register (NPR) captures diagnoses before 1997 using ICD-9 codes. The ICD-9 codes were converted from the ICD-10 codes using the conversion table from the Swedish National Board of Health and Welfare [6]. In the geriatric inpatient care cohort, 3-digit ICD-10 codes were used, as 4-digit ICD-10 codes were not available. In SveDem, 4-digit ICD-10 codes were used to identify chronic diseases from the NPR, and ATC codes were additionally used to identify further cases from the National Prescribed Drug Register, addressing the limitation that NPR only captures diagnoses from specialist care settings. The ICD codes and ATC codes corresponding to each chronic disease are shown in **Table 4**.

**Table 4. ICD codes and ATC codes for chronic diseases**

| Chronic disease | ICD-9 codes | ICD-10 codes, 3-digit or 4-digit | ICD-10 codes, 3-digit | ATC codes |
|---|---|---|---|---|
| Ischemic heart disease | 410,411,412,413,4140,4141,4148,4149, 4169,4292,V434 | I20,I21,I22,I24,I25,Z951,Z955 | I20,I21,I22,I23,I24,I2 5 | C01DA,C01EB18 |
| Heart failure | 4020,4021,4029,4040,4041,4049,4210, 4250,4251,4252,4253,4254,4255,4259, 4280,4281,4289,4291,4299,V421,V428 | I110,I130,I132,I27,I280,I42,I43, I50,I515,I517,I528,Z941,Z943 | I50 | C01A,C01CA02,C01CA0 7,C01CX08,C09DX04,C0 1EB17 |
| Stroke | 430,431,435,436,438,3448,3526,4320,4 321,4329,4330,4331,4332,4333,4338,4 339,4340,4341,4349,4370,4371,4372,4 373,4374,4375,4376,4378,4379 | G45,G46,I60,I61,I62,I63,I64,I67 ,I69 | I60,I61,I62,I63 | |
| Parkinson’s disease | 7360,2941,3320 | G20 | G20 | N04B |
| Chronic obstructive pulmonary disease | 4910,4911,4918,4789,4919,492,496,49 12,494 | J41,J42,J43,J44,J47 | J44 | |
| Rheumatoid arthritis | 7140,7148 | M059,M060 | M06 | |
| Osteoporosis | 7331,7330,7337,7308A | M80,M81,M82 | M80,M81 | |
| Cancer | 1400,1401,1403,1404,1405,1406,1408, 1409,1410,1411,1412,1413,1414,1415, 1416,1418,1419,1420,1421,1422,1428, 1429,1430,1431,1438,1439,1440,1441, 1448,1449,1450,1451,1452,1453,1454, 1455,1456,1458,1459,1460,1461,1462, 1463,1464,1465,1466,1467,1468,1469, 1470,1471,1472,1473,1478,1479,1480, 1481,1482,1483,1488,1489,1490,1491, | C00–C97 | C00-C97 | L01,L02BA |

| | |
|---|---|
| | 1498,1500,1501,1502,1503,1504,1505,1508,1509,1510,1511,1512,1513,1514,1515,1516,1518,1519,1520,1521,1522,1523,1528,1529,1530,1531,1532,1533,1534,1535,1536,1537,1538,1539,1540,1541,1542,1543,1548,1550,1551,1552,1560,1561,1562,1568,1569,1570,1571,1572,1573,1574,1578,1579,1580,1588,1589,1590,1591,1598,1599,1600,1601,1602,1603,1604,1605,1608,1609,1610,1611,1612,1613,1618,1619,1620,1622,1623,1624,1625,1628,1629,1630,1631,1638,1639,1640,1641,1642,1643,1648,1649,1650,1658,1659,1700,1701,1702,1703,1704,1705,1706,1707,1708,1709,1710,1712,1713,1714,1715,1716,1717,1718,1719,1720,1721,1722,1723,1724,1725,1726,1727,1728,1729,1730,1731,1732,1733,1734,1735,1736,1737,1738,1739,1740,1741,1742,1743,1744,1745,1746,1748,1749,175,179,1800,1801,1808,1809,181,1820,1821,1828,1830,1832,1833,1834,1835,1838,1839,1840,1841,1842,1843,1844,1848,1849,185,1860,1869,1871,1872,1873,1874,1875,1876,1877,1878,1879,1880,1881,1882,1883,1884,1885,1886,1887,1888,1889,1890,1891,1892,1893,1894,1898,1899,1900,1901,1902,1903,1904,1905,1906,1907,1908,1909,1910,1911,1912,1913,1914,1915,1916,1917,1918,1919,1920,1921,1922,1923,1928,1929,193,1940,1941,1943,1944,1945,1946,1948,1949,1950,1951,1952,1953,1954,1955,1958,1960,1961,1962,1963,1965,1966,1968,1969,1 |

| | | | | |
|---|---|---|---|---|
| | 970,1971,1972,1973,1974,1975,1976,1977,1978,1980,1981,1982,1983,1984,1985,1986,1987,1988,1990,1991,2000,2001,2002,2008,2010,2011,2012,2014,2015,2016,2017,2019,2020,2021,2022,2023,2024,2025,2026,2028,2029,2030,2031,2038,2040,2041,2042,2048,2049,2050,2051,2052,2053,2058,2059,2060,2061,2062,2068,2069,2070,2071,2072,2078,2080,2081,2082,2088,2089,2362,2386,2387,2389,2732,2733,2898,7573 | | | |
| Depression | 2958,2963,3004,2980,2961,2969,311,2966,3011,3090,3091,3092,3093,3094,3098,3099 | F204,F313,F314,F315,F32,F33,F341,F412,F432 | F32,F33 | N06A |
| Dementia | 2900,2901,2902,2904,2908,2909,2941,2953,3101,2903,3310,3311,3312,303,3498,3480,3368,3348,3342,3308,3349,3319,3309,3489 | F00,F01,F02,F03,F051,G30,G31 | F00,F01,F02,F03,G30 | |

Abbreviations: ATC = Anatomical Therapeutic Chemical Classification System, ICD-9 = the International Classification of Diseases, Ninth Revision, ICD-10 = the International Classification of Diseases, Tenth Revision.

### 4. Estimation of the number of older adults with malnutrition and healthcare costs of malnutrition

The number of older adults with malnutrition in Sweden in 2024 was estimated using (1) population statistics from Statistics Sweden (SCB), (2) data on the number of individuals admitted to geriatric inpatient care provided by the Swedish National Board of Health and Welfare (Socialstyrelsen), and (3) prevalences of malnutrition estimated in the current study (**Table 5**). The prevalences of malnutrition estimated from the SNAC-K were used as proxies for malnutrition among older adults not admitted to geriatric inpatient care, whereas the prevalences of malnutrition estimated from the geriatric inpatient care were used as proxies for those admitted to geriatric inpatient care.

The care costs of malnutrition in Sweden in 2024 were then estimated by multiplying the number of people with malnutrition by the costs of malnutrition per person in each age group and population and summing across all groups. For older adults admitted to geriatric inpatient care, costs were estimated using data from the geriatric inpatient care cohort, which included only 6 months of healthcare resource use. Annual costs for this group were extrapolated by assuming similar resource use in the latter 6 months of the year, while excluding costs associated with the initial geriatric inpatient admission.

**Table 5. Statistics for calculating the costs of malnutrition in Sweden at the national level**

| Population | Statistics | Source | Age group | | | | | | |
|---|---|---|---|---|---|---|---|---|---|
| | | | **60-64** | **65-69** | **70-74** | **75-79** | **80-84** | **85-89** | **90+** |
| Inhabitants in Sweden in 2024 | Number of inhabitants | SCB | 608482 | 548406 | 507193 | 503243 | 354101 | 189017 | 104528 |
| People admitted to geriatric inpatient care in Sweden in 2024 | Number of people | Socialstyrelsen | 978 | 2376 | 4759 | 9402 | 11604 | 10870 | 9588 |
| Older adults admitted to geriatric inpatient care | Prevalence of being at risk of malnutrition | The current study | 54.7% | 54.7% | 53.1% | 53.1% | 55% | 55% | 55% |
| | Prevalence of malnutrition | | 22.8% | 22.8% | 23.7% | 23.7% | 25.7% | 25.7% | 25.7% |
| Older adults not admitted to geriatric inpatient care | Prevalence of being at risk of malnutrition | | 23.3% | 23.3% | 23.3% | 34.9% | 34.9% | 34.9% | 34.9% |
| | Prevalence of malnutrition | | 0.8% | 0.8% | 0.8% | 6.9% | 6.9% | 6.9% | 6.9% |
| Older adults admitted to geriatric inpatient care | Annual costs of being at risk of malnutrition per person (2024 SEK) | | 117217 | 117217 | 117217 | 117217 | 117217 | 117217 | 117217 |
| | Annual costs of malnutrition per person (2024 SEK) | | 165470 | 165470 | 165470 | 165470 | 165470 | 165470 | 165470 |

| | | | | | | | | | |
|---|---|---|---|---|---|---|---|---|---|
| Older adults not admitted to geriatric inpatient care | Annual costs of being at risk of malnutrition per person (2024 SEK) | | 2267 | 2267 | 2267 | 2267 | 2267 | 2267 | 2267 |
| | Annual costs of malnutrition per person (2024 SEK) | | 1846 | 1846 | 1846 | 1846 | 1846 | 1846 | 1846 |

Abbreviation: SEK = Swedish kronor.
