## Supplementary tables for "Malnutrition and healthcare costs in older adults in Sweden: a longitudinal study based on a population-based cohort and Swedish registers"

**Table S1. Baseline characteristics of the SNAC-K study population**

| | Age group | | Overall (N=2982) |
|---|---|---|---|
| | 60-72 years (N=1553) | 78+ years (N=1429) | |
| **Age (years), mean (SD)** | 65.6 (4.9) | 85.2 (6.2) | 75.0 (11.3) |
| **Female sex, n (%)** | 893 (57.5) | 1044 (73.1) | 1937 (65.0) |
| **Education, n (%)** | | | |
| Elementary | 127 (8.2) | 393 (27.5) | 520 (17.4) |
| High school | 713 (45.9) | 757 (53.0) | 1470 (49.3) |
| University | 713 (45.9) | 255 (17.8) | 968 (32.5) |
| Missing | 0 (0) | 24 (1.7) | 24 (0.8) |
| **Living alone, n (%)** | 695 (44.8) | 1060 (74.2) | 1755 (58.9) |
| **Smoking status, n (%)** | | | |
| Never | 599 (38.6) | 639 (44.7) | 1238 (41.5) |
| Former | 644 (41.5) | 396 (27.7) | 1040 (34.9) |
| Current | 278 (17.9) | 105 (7.3) | 383 (12.8) |
| Missing | 32 (2.1) | 289 (20.2) | 321 (10.8) |
| **Nutritional status by MNA definition, n (%)** | | | |
| Normal nutritional status | 1147 (73.9) | 771 (54.0) | 1918 (64.3) |
| At risk of malnutrition | 351 (22.6) | 430 (30.1) | 781 (26.2) |
| Malnourished | 12 (0.8) | 80 (5.6) | 92 (3.1) |
| Missing | 43 (2.8) | 148 (10.4) | 191 (6.4) |
| **Chronic diseases, n (%)** | | | |
| Ischemic heart disease | 154 (9.9) | 360 (25.2) | 514 (17.2) |
| Heart failure | 45 (2.9) | 309 (21.6) | 354 (11.9) |
| Stroke | 79 (5.1) | 226 (15.8) | 305 (10.2) |
| Parkinson's disease | 11 (0.7) | 20 (1.4) | 31 (1.0) |
| Chronic obstructive pulmonary disease | 68 (4.4) | 89 (6.2) | 157 (5.3) |
| Rheumatoid arthritis | 19 (1.2) | 40 (2.8) | 59 (2.0) |
| Osteoporosis | 60 (3.9) | 172 (12.0) | 232 (7.8) |
| Cancer | 266 (17.1) | 337 (23.6) | 603 (20.2) |
| Depression | 150 (9.7) | 166 (11.6) | 316 (10.6) |
| Dementia | 18 (1.2) | 146 (10.2) | 164 (5.5) |
| **Loss of smell, n (%)** | 270 (17.4) | 316 (22.1) | 586 (19.7) |
| Missing | 15 (1.0) | 111 (7.8) | 126 (4.2) |

| | Age group | | Overall (N=2982) |
|---|---|---|---|
| | 60-72 years (N=1553) | 78+ years (N=1429) | |
| **Loss of taste, n (%)** | 125 (8.0) | 199 (13.9) | 324 (10.9) |
| Missing | 14 (0.9) | 100 (7.0) | 114 (3.8) |
| **Difficulty with chewing food, n (%)** | 133 (8.6) | 341 (23.9) | 474 (15.9) |
| Missing | 7 (0.5) | 60 (4.2) | 67 (2.2) |
| **Reduced appetite, n (%)** | 66 (4.2) | 254 (17.8) | 320 (10.7) |
| Missing | 12 (0.8) | 104 (7.3) | 116 (3.9) |

Abbreviations: MNA = Mini-Nutritional Assessment, SD = standard deviation, SNAC-K = The Swedish National Study on Aging and Care in Kungsholmen.

**Table S2. Baseline characteristics of the geriatric inpatient care cohort**

| | Age group | | | Overall (N=7680) |
|---|---|---|---|---|
| | <70 (N=449) | 70-79 (N=2027) | 80+ (N=5204) | |
| **Age (years), mean (SD)** | 65.2 (3.9) | 75.1 (2.7) | 87.8 (5.0) | 83.1 (8.4) |
| **Female sex, n (%)** | 245 (54.6) | 1129 (55.7) | 3319 (63.8) | 4693 (61.1) |
| **Education, n (%)** | | | | |
| Primary school | 19 (4.2) | 249 (12.3) | 901 (17.3) | 1169 (15.2) |
| Lower secondary school | 95 (21.2) | 249 (12.3) | 488 (9.4) | 832 (10.8) |
| Upper secondary school | 176 (39.2) | 663 (32.7) | 1488 (28.6) | 2327 (30.3) |
| Post-secondary school | 83 (18.5) | 416 (20.5) | 904 (17.4) | 1403 (18.3) |
| Higher than post-secondary school | 24 (5.3) | 193 (9.5) | 399 (7.7) | 616 (8.0) |
| Missing | 52 (11.6) | 257 (12.7) | 1024 (19.7) | 1333 (17.4) |
| **Living alone, n (%)** | 239 (53.2) | 1035 (51.1) | 3235 (62.2) | 4509 (58.7) |
| Missing | 12 (2.7) | 19 (0.9) | 11 (0.2) | 42 (0.5) |
| **Nutritional status by MNA definition, n (%)** | | | | |
| Normal nutritional status | 95 (21.2) | 440 (21.7) | 943 (18.1) | 1478 (19.2) |
| At risk of malnutrition | 233 (51.9) | 1002 (49.4) | 2679 (51.5) | 3914 (51.0) |
| Malnourished | 98 (21.8) | 447 (22.1) | 1251 (24.0) | 1796 (23.4) |
| Missing | 23 (5.1) | 138 (6.8) | 331 (6.4) | 492 (6.4) |
| **Functional ability, n (%)** | | | | |
| Independent | 14 (3.1) | 60 (3.0) | 84 (1.6) | 158 (2.1) |

| | Age group | | | Overall (N=7680) |
|---|---|---|---|---|
| | <70 (N=449) | 70-79 (N=2027) | 80+ (N=5204) | |
| Slightly dependent | 19 (4.2) | 68 (3.4) | 94 (1.8) | 181 (2.4) |
| Moderately dependent | 146 (32.5) | 723 (35.7) | 1617 (31.1) | 2486 (32.4) |
| Severely dependent | 155 (34.5) | 723 (35.7) | 2121 (40.8) | 2999 (39.0) |
| Totally dependent | 46 (10.2) | 207 (10.2) | 681 (13.1) | 934 (12.2) |
| Missing | 69 (15.4) | 246 (12.1) | 607 (11.7) | 922 (12.0) |
| **Chronic diseases, n (%)** | | | | |
| Ischemic heart disease | 52 (11.6) | 299 (14.8) | 989 (19.0) | 1340 (17.4) |
| Heart failure | 65 (14.5) | 384 (18.9) | 1566 (30.1) | 2015 (26.2) |
| Stroke | 49 (10.9) | 180 (8.9) | 417 (8.0) | 646 (8.4) |
| Parkinson's disease | 25 (5.6) | 107 (5.3) | 129 (2.5) | 261 (3.4) |
| Chronic obstructive pulmonary disease | 75 (16.7) | 433 (21.4) | 678 (13.0) | 1186 (15.4) |
| Rheumatoid arthritis | 9 (2.0) | 54 (2.7) | 93 (1.8) | 156 (2.0) |
| Osteoporosis | 36 (8.0) | 215 (10.6) | 687 (13.2) | 938 (12.2) |
| Cancer | 56 (12.5) | 314 (15.5) | 821 (15.8) | 1191 (15.5) |
| Depression | 35 (7.8) | 170 (8.4) | 346 (6.6) | 551 (7.2) |
| Dementia | 18 (4.0) | 210 (10.4) | 772 (14.8) | 1000 (13.0) |
| **Main cause of inpatient care admission, n (%)** | | | | |
| Heart failure | 11 (2.4) | 70 (3.5) | 392 (7.5) | 473 (6.2) |
| Fracture of femur | 31 (6.9) | 114 (5.6) | 291 (5.6) | 436 (5.7) |

| | Age group | | | Overall (N=7680) |
|---|---|---|---|---|
| | <70 (N=449) | 70-79 (N=2027) | 80+ (N=5204) | |
| Bacterial pneumonia | 11 (2.4) | 93 (4.6) | 186 (3.6) | 290 (3.8) |
| Cerebral infarction | 12 (2.7) | 74 (3.7) | 179 (3.4) | 265 (3.5) |
| Other chronic obstructive pulmonary disease | 13 (2.9) | 96 (4.7) | 115 (2.2) | 224 (2.9) |
| Osteoporosis with current pathological fracture | 16 (3.6) | 59 (2.9) | 144 (2.8) | 219 (2.9) |
| Disorders of urinary system | 9 (2) | 38 (1.9) | 168 (3.2) | 215 (2.8) |
| Fracture of lumbar spine and pelvis | 9 (2) | 45 (2.2) | 114 (2.2) | 168 (2.2) |
| Spondylopathies | 6 (1.3) | 55 (2.7) | 105 (2) | 166 (2.2) |
| Acute pyelonephritis | 8 (1.8) | 36 (1.8) | 115 (2.2) | 159 (2.1) |

Abbreviations: MNA = Mini-Nutritional Assessment, SD = standard deviation.

**Table S3. Baseline characteristics of the SveDem population**

| | Age group | | | Overall (N=64192) |
|---|---|---|---|---|
| | <70 (N=5871) | 70-79 (N=21658) | 80+ (N=36663) | |
| **Age (years), mean (SD)** | 64.2 (5.0) | 75.3 (2.7) | 85.6 (4.0) | 80.2 (7.9) |
| **Female sex, n (%)** | 2989 (50.9) | 11496 (53.1) | 22544 (61.5) | 37029 (57.7) |
| **Malnutrition, n (%)** | | | | |
| No | 3889 (66.2) | 12173 (56.2) | 17848 (48.7) | 33910 (52.8) |
| Yes | 419 (7.1) | 3561 (16.4) | 6455 (17.6) | 10435 (16.3) |
| Missing | 1563 (26.6) | 5924 (27.4) | 12360 (33.7) | 19847 (30.9) |
| **Dementia subtypes, n (%)** | | | | |
| Unspecified | 824 (14.0) | 3749 (17.3) | 8795 (24.0) | 13368 (20.8) |
| VaD | 743 (12.7) | 3651 (16.9) | 8099 (22.1) | 12493 (19.5) |
| Mixed AD/VaD | 453 (7.7) | 3789 (17.5) | 7965 (21.7) | 12207 (19.0) |
| Late-onset AD | 1043 (17.8) | 7950 (36.7) | 9901 (27.0) | 18894 (29.4) |
| Early-onset AD | 1655 (28.2) | 193 (0.9) | 87 (0.2) | 1935 (3.0) |
| PDD | 141 (2.4) | 513 (2.4) | 317 (0.9) | 971 (1.5) |
| FTD | 403 (6.9) | 429 (2.0) | 190 (0.5) | 1022 (1.6) |
| LBD | 193 (3.3) | 727 (3.4) | 515 (1.4) | 1435 (2.2) |
| MCI | 36 (0.6) | 51 (0.2) | 32 (0.1) | 119 (0.2) |
| Other | 380 (6.5) | 606 (2.8) | 762 (2.1) | 1748 (2.7) |
| **Living alone, n (%)** | 1835 (31.3) | 7421 (34.3) | 18862 (51.4) | 28118 (43.8) |
| Missing | 287 (4.9) | 786 (3.6) | 2810 (7.7) | 3883 (6.0) |
| **Institutionalized, n (%)** | 442 (7.6) | 1105 (5.1) | 3836 (10.5) | 5383 (8.4) |
| Missing | 17 (0.3) | 49 (0.2) | 65 (0.2) | 131 (0.2) |
| **Chronic diseases, n (%)** | | | | |
| Ischemic heart disease | 437 (7.4) | 2908 (13.4) | 7497 (20.4) | 10842 (16.9) |
| Heart failure | 206 (3.5) | 1687 (7.8) | 5360 (14.6) | 7253 (11.3) |
| Stroke | 645 (11.0) | 3093 (14.3) | 5713 (15.6) | 9451 (14.7) |
| Parkinson's disease | 308 (5.2) | 1213 (5.6) | 1127 (3.1) | 2648 (4.1) |
| Chronic obstructive pulmonary disease | 243 (4.1) | 1188 (5.5) | 1841 (5.0) | 3272 (5.1) |
| Rheumatoid arthritis | 70 (1.2) | 370 (1.7) | 563 (1.5) | 1003 (1.6) |
| Osteoporosis | 95 (1.6) | 795 (3.7) | 2383 (6.5) | 3273 (5.1) |
| Cancer | 518 (8.8) | 3276 (15.1) | 6875 (18.8) | 10669 (16.6) |

| | Age group | | | Overall (N=64192) |
|---|---|---|---|---|
| | <70 (N=5871) | 70-79 (N=21658) | 80+ (N=36663) | |
| Depression | 2046 (34.8) | 6559 (30.3) | 9611 (26.2) | 18216 (28.4) |

Abbreviations: AD = Alzheimer's disease, FTD = Frontotemporal dementia, LBD = Lewy Body dementia, MCI = Mild cognitive impairment, PDD = Parkinson's disease dementia, SD = standard deviation, SveDem = the Swedish Register of Cognitive/Dementia Disorders, VaD = Vascular dementia.

**Table S4. The association between malnutrition and annual healthcare costs in 2024 SEK in the SNAC-K**

| | Model 1 | | Model 2 | |
|---|---|---|---|---|
| | **Costs (95% CI)** | **Difference (95% CI)** | **Costs (95% CI)** | **Difference (95% CI)** |
| **Total costs** | | | | |
| Normal | 26724 (25539,27909) | Reference | 27575 (26365,28784) | Reference |
| At risk | 31706 (29473,33940) | 4982 (2543,7421) | 29841 (27841,31841) | 2267 (64,4469) |
| Malnourished | 33121 (25439,40803) | 6397 (-1391,14185) | 29420 (20985,37855) | 1846 (-6802,10493) |
| **Primary care costs** | | | | |
| Normal | 8281 (7905,8657) | Reference | 8564 (8178,8951) | Reference |
| At risk | 9815 (9115,10514) | 1533 (757,2310) | 9157 (8530,9785) | 593 (-127,1313) |
| Malnourished | 9395 (6394,12396) | 1114 (-1924,4151) | 7855 (4730,10981) | -709 (-3902,2483) |
| **Specialist outpatient care costs** | | | | |
| Normal | 4240 (4053,4427) | Reference | 4311 (4121,4500) | Reference |
| At risk | 5090 (4697,5483) | 850 (415,1286) | 4876 (4488,5263) | 565 (125,1005) |
| Malnourished | 5135 (3475,6794) | 895 (-774,2563) | 4906 (3373,6440) | 595 (-954,2145) |
| **Specialist inpatient care costs** | | | | |
| Normal | 17707 (16685,18729) | Reference | 18291 (17226,19356) | Reference |
| At risk | 20623 (18698,22548) | 2916 (722,5110) | 19201 (17417,20984) | 910 (-1209,3029) |
| Malnourished | 19050 (12642,25457) | 1343 (-5204,7890) | 18289 (12104,24474) | -2 (-6365,6361) |

Abbreviations: CI = confidence interval, SEK = Swedish kronor, SNAC-K = The Swedish National Study on Aging and Care in Kungsholmen.

Notes: Results are from marginal predictions from two-part models. Model 1 adjusted for age, sex, follow-up year, and death during the year. Model 2 additionally adjusted for smoking status, living alone, ischemic heart disease, heart failure, rheumatoid arthritis or osteoporosis, cancer, depression, and Parkinson’s disease or dementia.

**Table S5. The association between malnutrition and 6-month healthcare costs in 2024 SEK in the geriatric inpatient care cohort**

| | Model 1 | | Model 2 | |
|---|---|---|---|---|
| | Costs (95% CI) | Difference (95% CI) | Costs (95% CI) | Difference (95% CI) |
| **Total costs[a]** | | | | |
| Normal | 313332 (302259,324404) | Reference | 341558 (329385,353730) | Reference |
| At risk | 399471 (391061,407881) | 86139 (72066,100212) | 401763 (393335,410191) | 60205 (45613,74798) |
| Malnourished | 458522 (444299,472744) | 145190 (127056,163324) | 428176 (414979,441373) | 86619 (68362,104875) |
| **Care costs during stay at the geriatric inpatient care – first admission[a]** | | | | |
| Normal | 59583 (58692,60474) | Reference | 60809 (59902,61716) | Reference |
| At risk | 63884 (63303,64466) | 4301 (3229,5374) | 64002 (63433,64572) | 3194 (2121,4267) |
| Malnourished | 69975 (69027,70923) | 10392 (9084,11701) | 68575 (67636,69513) | 7766 (6419,9114) |
| **Care costs during stay at the geriatric inpatient care – readmissions[b]** | | | | |
| Normal | 18538 (16310,20766) | Reference | 18955 (16662,21247) | Reference |
| At risk | 21917 (20463,23371) | 3379 (671,6086) | 21771 (20333,23210) | 2817 (76,5558) |
| Malnourished | 21709 (19565,23853) | 3171 (82,6259) | 21655 (19454,23855) | 2700 (-548,5948) |
| **Primary care costs[b]** | | | | |
| Normal | 61105 (56528,65682) | Reference | 64248 (59302,69194) | Reference |
| At risk | 72111 (68778,75444) | 11007 (5273,16740) | 72422 (69021,75823) | 8174 (2267,14082) |
| Malnourished | 69986 (65016,74956) | 8881 (2135,15628) | 68590 (63683,73497) | 4342 (-2695,11379) |
| **Specialist outpatient care costs[b]** | | | | |
| Normal | 25420 (24017,26823) | Reference | 24678 (23367,25988) | Reference |
| At risk | 27026 (26088,27964) | 1606 (-95,3307) | 26617 (25739,27496) | 1940 (371,3509) |
| Malnourished | 24284 (23003,25566) | -1135 (-3025,754) | 25562 (24232,26891) | 884 (-1011,2779) |
| **Other specialist inpatient care costs[b]** | | | | |
| Normal | 43694 (39536,47852) | Reference | 43381 (39238,47524) | Reference |
| At risk | 51325 (48545,54104) | 7631 (2557,12705) | 51004 (48272,53736) | 7623 (2602,12644) |
| Malnourished | 50897 (46907,54888) | 7204 (1426,12981) | 51906 (47725,56087) | 8525 (2476,14574) |
| **Social care costs[b]** | | | | |

| | | | | |
|---|---|---|---|---|
| Normal | 107222 (100208,114237) | Reference | 132825 (124571,141080) | Reference |
| At risk | 166934 (161419,172449) | 59712 (50746,68677) | 170383 (164774,175993) | 37558 (27945,47171) |
| Malnourished | 223891 (213803,233979) | 116669 (104215,129122) | 192286 (183330,201242) | 59461 (46937,71984) |

Abbreviations: CI = confidence interval, SEK = Swedish kronor.
Notes:
[a]Results are from marginal predictions from generalized linear regression models. Model 1 adjusted for age and sex. Model 2 additionally adjusted for civil status, educational level, living alone, functional ability, heart failure, depression, Parkinson’s disease, and dementia.
[b]Results are from marginal predictions from two-part models. Model 1 adjusted for age and sex. Model 2 additionally adjusted for civil status, educational level, functional ability, living alone, heart failure, depression, Parkinson’s disease, and dementia.

**Table S6. The association between malnutrition and annual healthcare costs in 2024 SEK in SveDem**

| | Model 1 | | Model 2 | |
|---|---|---|---|---|
| | Costs (95% CI) | Difference (95% CI) | Costs (95% CI) | Difference (95% CI) |
| **Total costs** | | | | |
| Normal | 501084 (497927,504242) | Reference | 505276 (502354,508199) | Reference |
| Malnourished | 541513 (535144,547883) | 40429 (32952,47906) | 527446 (521489,533404) | 22170 (15152,29188) |
| **Prescribed drug costs** | | | | |
| Normal | 10482 (10412,10553) | Reference | 10504 (10438,10571) | Reference |
| Malnourished | 9802 (9660,9944) | -680 (-850,-510) | 9730 (9600,9861) | -774 (-929,-618) |
| **Specialist outpatient care costs** | | | | |
| Normal | 11911 (11820,12001) | Reference | 11869 (11782,11955) | Reference |
| Malnourished | 11567 (11379,11755) | -343 (-567,-119) | 11710 (11532,11887) | -159 (-368,51) |
| **Specialist inpatient care costs** | | | | |
| Normal | 45497 (44978,46017) | Reference | 45458 (44945,45971) | Reference |
| Malnourished | 45175 (44168,46182) | -322 (-1513,870) | 45304 (44305,46303) | -154 (-1337,1030) |
| **Social care costs** | | | | |
| Normal | 428261 (425391,431130) | Reference | 432769 (430146,435392) | Reference |
| Malnourished | 470821 (465001,476642) | 42561 (35716,49405) | 455463 (450178,460748) | 22694 (16487,28901) |

Abbreviations: CI = confidence interval, SEK = Swedish kronor, SveDem = the Swedish Register of Cognitive/Dementia Disorders.

Notes: Results are from marginal predictions from two-part models. Model 1 adjusted for age, sex, dementia subtypes, follow-up year, and death during the year. Model 2 additionally adjusted for institutionalization status, living alone, severity of cognitive impairment, ischemic heart disease, heart failure, stroke, chronic obstructive pulmonary disease, rheumatoid arthritis, osteoporosis, and cancer.

**Table S7. Estimated costs of malnutrition in Swedish older adults in 2024 at the national level**

| | At risk | Malnourished | Total |
|---|---|---|---|
| **Total number of individuals** | 800000 | 102000 | 902000 |
| Individuals admitted to geriatric inpatient care | 27000 | 12000 | 39000 |
| Individuals not admitted to geriatric inpatient care | 773000 | 90000 | 863000 |
| **Costs associated with malnutrition per person (2024 SEK)** | | | |
| Individuals admitted to geriatric inpatient care | 117217 | 165470 | |
| Individuals not admitted to geriatric inpatient care | 2266 | 1845 | |
| **Total costs (2024 SEK million)** | 4916 | 2211 | 7127 |
| Individuals admitted to geriatric inpatient care | 3164 | 2045 | 5208 |
| Individuals not admitted to geriatric inpatient care | 1752 | 166 | 1918 |

Abbreviation: SEK = Swedish kronor.
